# Multiplexed FRET-FLIM Profiling of Immune Checkpoint Interactions Predicts Response to Atezolizumab in Urothelial Carcinoma

**DOI:** 10.64898/2026.09.01.26361904

**Authors:** Laura Camacho, Cristina Cacho-Navas, Jon Agüero, Baterdene Batmunkh, José María Gracia, Killian O’ Sullivan, Markel Rementeria, James Miles, Juan Gumuzio, Fernando Aguirre, Salvador Martin Algarra, Carlos Eduardo de Andrea, Peter J. Parker, Véronique Calleja

**Affiliations:** HAWK Biosystems (FASTBASE Solutions S.L.) 612 Astondo Bidea, Science and Technology Park of Bizkaia, 48160 Derio, Spain; Facultad de Medicina, Departamento de Oncología, Clínica Universidad de Navarra (CUN), 31008 Pamplona, Spain; Facultad de Medicina, Departamento de Patología, Anatomía y Fisiología, Clínica Universidad de Navarra (CUN), 31008 Pamplona, Spain; School of Cancer and Pharmaceutical Sciences, Comprehensive Cancer Centre, King’s College London, SE1 1UL, London, United Kingdom; Francis Crick Institute, NW1 1AT, London, United Kingdom

**Keywords:** Functional Proteomics, Spatial Biology, QF-Pro, FRET-FLIM, Immune Checkpoint Therapy, Precision Medicine, Urothelial carcinoma

## Abstract

Immune checkpoint inhibitors targeting the PD-1/PD-L1 axis have shown great promise in treating bladder cancer and are now part of the standard treatment for advanced disease. However, many patients still fail to respond to treatment and at present many biomarkers are assessed but have yet shown only limited results. Therefore, with the advent of combination treatments and the increase of immune related adverse event, the search for reliable predictive biomarkers is paramount. Using a multiplexed enhanced FRET-FLIM based technique (QF-Pro) we quantified the interaction of PD-1/PD-L1, CTLA-4/CD80 and TIGIT/CD155 immune checkpoints in a pre-treatment TMA of 46 patients treated with atezolizumab. The association between higher PD-1/PD-L1 ICP interaction state and treatment efficacy was demonstrated in the male sample cohort, where it identified patients with better PFS. Conversely, patients exhibiting higher CTLA-4/CD80 engagement had a worse response to atezolizumab. Remarkably, the dual assessment of patients with high PD-1/PD-L1 and low CTLA-4/CD80 allowed to identify the best responders. These results indicate that the monitoring of patients’ immune profile in urothelial carcinoma might be critical in identifying patients who may benefit from combination therapy.

## Introduction

Urothelial carcinoma is the most prevalent malignancy of the urinary tract, with a markedly worse prognosis in muscle-invasive and metastatic disease [1]. The approval of immune checkpoint inhibitors (ICIs) targeting PD-1/PD-L1 axis, including pembrolizumab (anti-PD-1) and atezolizumab (anti-PD-L1) whose approval was supported by the IMvigor210 trial for the treatment of advanced urothelial carcinoma (UC) [2, 3], have transformed the treatment of cisplatin-ineligible patients with advanced urothelial cancer [4]. Unfortunately, single-agent anti-PD-L1 therapy yields objective responses in only 20 to 25% of patients [5]. More recently, targeted therapy using the new antibody-drug conjugate enfortumab vedotin, combined with pembrolizumab has become the preferred first-line regimen in eligible patients [6]. However, the growing adoption of combination ICI regimens carries heightened risk of immune-related adverse events (irAEs) [7, 8]. Additionally, beyond PD-1/PD-L1 checkpoint engagement, sustained immune suppression involves multiple co-inhibitory axes operating in parallel. Among these, the CTLA-4 axis plays a crucial role in immunosuppression by attenuating T cell priming and sustaining an immunosuppressive Treg-rich environment in many tumours, however its role in urothelial carcinoma trial has yielded inconclusive results (DANUBE trial; [8]). In muscle-invasive bladder cancer, elevated TIGIT expression on tumour-infiltrating T cells has been linked to worse clinical outcomes [9], and high expression of its ligand and specifically membranous CD155 was associated with poor prognosis in bladder cancer [10]. Furthermore, PD-1 blockade has been shown to upregulate TIGIT expression as a compensatory mechanism in adaptive immune resistance in urothelial as in other solid tumours, providing mechanistic rationale for a possible dual checkpoint targeting [11].

In view of these complexities, evaluating multiple immunosuppressive axes which would provide a more complete map of the tumour microenvironment compared to a single biomarker assessment should better guide the selection of therapeutic regimen.

Currently, the main biomarker strategy to inform on ICI treatment relies on immunohistochemistry with the use of PD-L1 expression scores (TPS/CPS). Yet across pivotal trials, PD-L1 expression has shown only inconsistent predictive value, confounded by inter-assay, scoring, and tissue compartment variability [12, 13]. While PD-L1 quantifies protein abundance it does not directly reflects the functional checkpoint engagement, the real target of the ICI treatment. With the expanding use of combination ICI regimens and the attendant risks of immune related adverse events (irAEs), the identification of robust predictive biomarkers that accurately reflect the functional immune state of the tumour microenvironment (TME) is an important clinical priority.

We previously showed that in non-small cell lung cancer (NSCLC), using QF-Pro, an amplified FRET-FLIM-based technology platform, that a high PD-1/PD-L1 interaction state, but not the conventional PD-L1 tumour proportion score (TPS), was predictive of patient response to anti-PD-1/PD-L1 blockade [14]. These results demonstrated the clinical superiority of interaction-level quantification over expression-based measurements. Here, we applied QF-Pro multiplexed FRET-FLIM to a FFPE TMA of pre-treatment tissue of patients obtained from a real-world cohort of 109 advanced urothelial cancer patients treated with atezolizumab [15]. A prior analysis of this TMA on n=45 patients had demonstrated that intratumoral CD8+ T cell density and co-expression of PD-1, TIM-3 and LAG-3 on CD8+ T cells were significantly associated with best overall response [15]. Building on this characterisation, we aimed to quantify the functional engagement states of PD-1/PD-L1, CTLA-4/CD80 and TIGIT/CD155, not yet evaluated in these tumours and hence assess their association with progression-free survival (PFS). We report that high PD-1/PD-L1 identify patients with superior PFS, while high CTLA-4/CD80 engagement is associated with reduced benefit from anti-PD-L1 monotherapy, a finding with direct implications for selecting patients who may benefit from dual anti-CTLA-4/anti-PD-L1 combination therapy.

## Results

### Multiplex quantification of three immune checkpoint interactions in a retrospective study cohort of urothelial carcinoma patients treated with atezolizumab

The retrospective urothelial cohort encompassed patients treated with atezolizumab (n=46) with evaluable pre-treatment formalin-fixed paraffin-embedded (FFPE) tumour tissue compatible with amplified FRET-FLIM analysis. The TMA included two cores per patient from different locations in the tumour. A comprehensive table of the cohort clinical characteristics can be found in Table 1. The clinical outcomes were assessed using overall best response to atezolizumab: complete response (CR), partial response (PR), stable disease (SD), progressive disease (PD) and progression free survival (PFS).

**Table 1.** Baseline clinical characteristics of atezolizumab-treated urothelial carcinoma patients

| Characteristic | All patients<br>(n = 46) | Males<br>(n = 34) | Females<br>(n = 12) |
| --- | --- | --- | --- |
| **Age** |  |  |  |
| Median, years (range) | 68 (48–82) | 69 (48–81) | 63 (51–82) |
| **Sex** |  |  |  |
| Male | 34 (73.9%) | - | - |
| Female | 12 (26.1%) | - | - |
| **Disease setting** |  |  |  |
| Primary | 37 (80.4%) | 28 (82.4%) | 9 (75.0%) |
| Metastatic | 5 (10.9%) | 3 (8.8%) | 2 (16.7%) |
| Not evaluable | 4 (8.7%) | 3 (8.8%) | 1 (8.3%) |
| **Prior lines of therapy** |  |  |  |
| 1 line | 20 (43.5%) | 16 (47.1%) | 4 (33.3%) |
| 2 lines | 11 (23.9%) | 7 (20.6%) | 4 (33.3%) |
| ≥3 lines | 8 (17.4%) | 7 (20.6%) | 1 (8.3%) |
| Missing data | 7 (15.2%) | 4 (11.8%) | 3 (25.0%) |
| **Best overall response (RECIST 1.1)** |  |  |  |
| Complete response (CR) | 5 (10.9%) | 3 (8.8%) | 2 (16.7%) |
| Partial response (PR) | 10 (21.7%) | 10 (29.4%) | 0 (0.0%) |
| Stable disease (SD) | 13 (28.3%) | 10 (29.4%) | 3 (25.0%) |
| Progression disease (PD) | 18 (39.1%) | 11 (32.4%) | 7 (58.3%) |
| **Objective response rate (ORR; CR+PR)** |  |  |  |
| n (%) | 15 (32.6%) | 13 (38.2%) | 2 (16.7%) |
| **Disease control rate (DCR; CR+PR+SD)** |  |  |  |
| n (%) | 28 (60.9%) | 23 (67.6%) | 5 (41.7%) |
| **Progression-free survival (PFS)** |  |  |  |
| Median, months (range) | 5.8 (0.3–20.4) | 6.0 (0.3–20.4) | 2.8 (0.9–18.7) |

The multiplexed quantification of PD-1/PD-L1, CTLA-4/CD80 and TIGIT/CD155 immune checkpoint (ICP) interactions (Fig. 1A) was performed on the full cohort (n = 46) using the QF-Pro technology platform and reagents (Supplementary Fig. 1 and Materials and Methods). In subsequent analysis the ICP interaction values obtained from the multiple fields within each of the two tumour cores per patient were combined, thereby incorporating intra-tumoral heterogeneity. The spatial localisation of the ICP proteins (green and red) and interaction (QF-Pro map; magenta) in one patient core is exemplified in figure 1B. The ICP interaction scores (QF-Pro scores) for this patient, are presented on the graph (Fig. 1B right panel). Each dot corresponds to a field of view from the core imaged.

**Figure 1.**
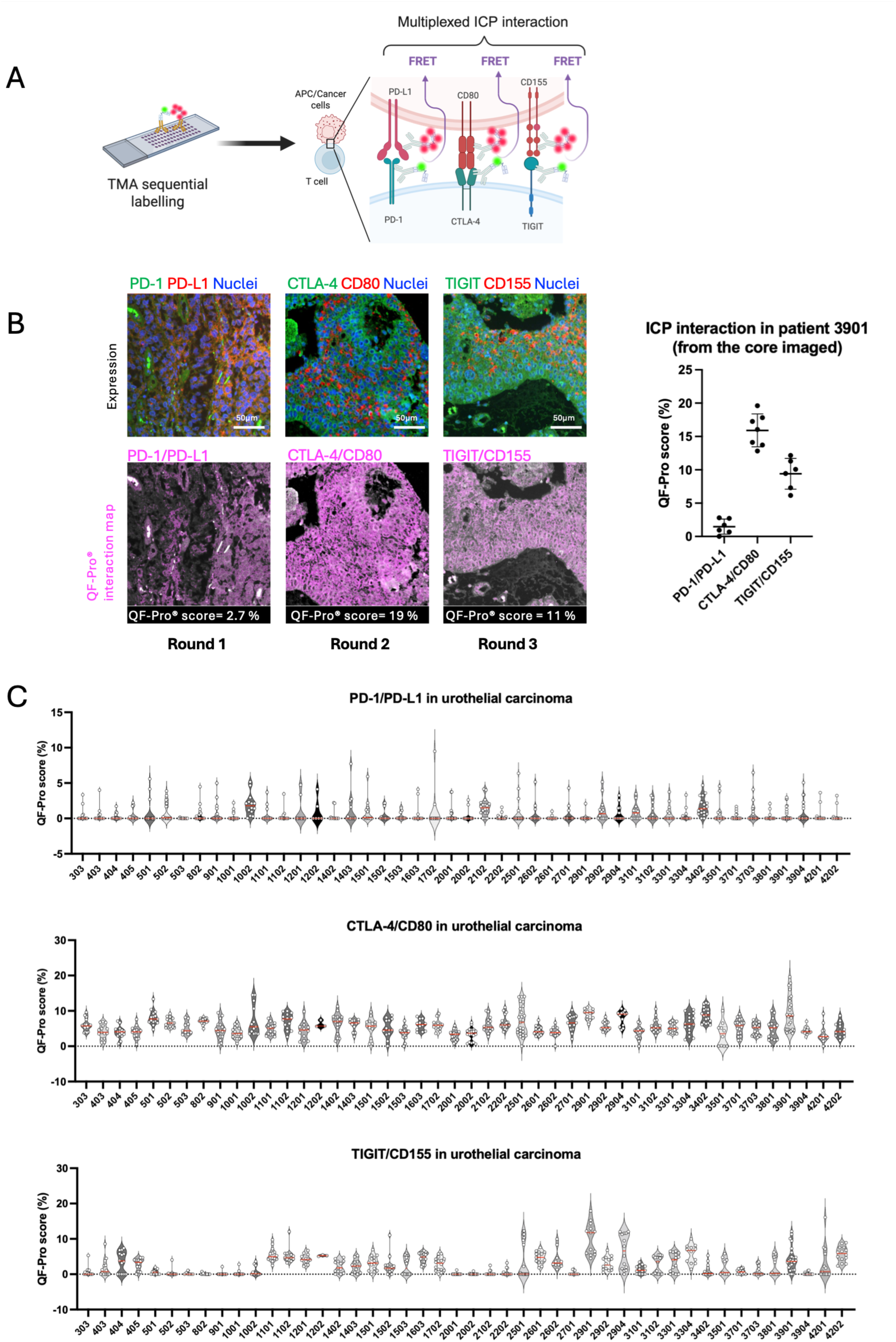
Multiplexed QF-Pro FRET-FLIM profiling of immune checkpoint interactions in FFPE urothelial carcinoma tissue. **(A)** Cartoon showing the three immune checkpoint interactions quantified in the urothelial TMA. Mouse and rabbit primary antibodies targeting the respective receptors and ligands are detected using anti-mouse F(ab’)₂–ATTO488 (donor, green) and anti-rabbit F(ab’)₂–HRP secondary antibodies. Signal amplification is achieved by tyramide labelling (acceptor, red), enabling detection of FRET for each immune checkpoint interaction. (**B**) Representative fluorescence images from three sequential rounds of multiplex staining in a tissue core (patient # 3901). Each round shows protein expression (green and red) and DAPI nuclear counterstain (blue) alongside the corresponding QF-Pro interaction map, in which pixel colour encodes the FRET-derived interaction scores (QF-Pro scores, magenta) for PD-1/PD-L1 (11%); CTLA-4/CD80 (19%) and TIGIT/CD155 (2.7%). The bar chart (right panel) shows the interaction scores for the three immune checkpoint protein (ICP) in the same patient #3901. Each dot represent a field of view of the core. **(C)** Violin plots of the interaction scores distribution for PD-1/PD-L1, CTLA-4/CD80, and TIGIT/CD155 across all 46 patients. Each dot represents one field of view.

The violin plots illustrate both intra-and inter-patients heterogeneity in the three ICP interactions scores for the same patients across the full cohort, with each dot representing a field of view from the cores (Fig. 1C).

### Independent interaction of the three immune checkpoints engagement in the urothelial cohort

The multiplexed profiling of the three ICP interactions allowed us to explore the potential exclusion or association between the interaction states of these checkpoints. The ICP interaction scores relationships were assessed by performing pairwise Pearson’s correlation analyses across the 46 patients (Fig. 2A). The correlation heatmap and corresponding scatter plot showing individual patient interaction scores (Fig. 2B) indicate weak correlations that do not reached statistical significance: PD-1/PD-L1 vs CTLA-4/CD80 (r=0.21, p=0.153), PD-1/PD-L1 vs TIGIT/CD155 (r=-0.06, p=0.696), and CTLA-4/CD80 vs TIGIT/CD155 (r=0.10, p=0.494). A patient (red dot) unusually displaying a very high value for TIGIT/CD155 interaction (∼10%) was identified as an outlier by both Z-score (|z|>2.5) and Turkey’s fence of 9.271 and was therefore excluded from the correlation analysis to avoid biasing the interpretation. The absence of significant inter-biomarker correlation indicated that the three checkpoint interactions were largely independently distributed across tumours in this cohort.

**Figure 2.**
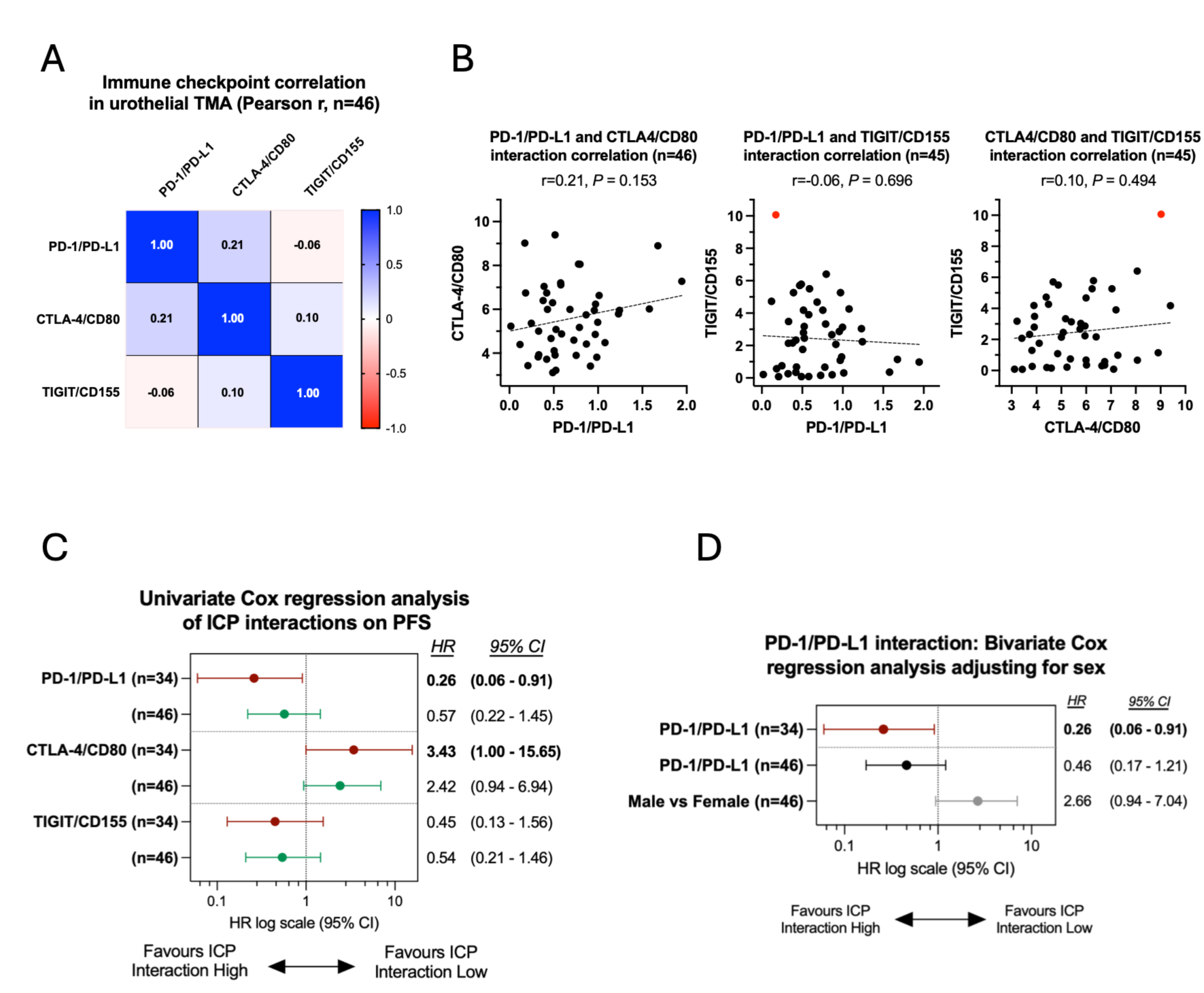
Immune checkpoint interactions correlation and association with PFS in atezolizumab-treated urothelial carcinoma patients. **(A)** Pearson’s correlation coefficients (r) for PD-1/PD-L1, CTLA-4/CD80, and TIGIT/CD155 interaction states for all evaluable patients (n = 46). The r coefficients are presented in a color-coded matrix, with red indicating negative correlations, blue indicating positive correlations, and colour intensity reflecting correlation strength. **(B)** Pairwise Pearson’s correlation scatter plots for continuous FRET interaction scores across all three ICP pairs. Each point represents one patient (n=46). Pearson r and two-tailed *P* values are shown above the graph. No significant correlation was observed between any pair. The red dot identified as an outlier using simultaneous Z-score >2.5 and Turkey’s fence, was excluded from the pairwise Pearson’s correlation calculations. **(C)** Univariate Cox proportional hazards model regression was performed to estimate hazard ratios (HR) and 95% confidence intervals (CI) for high vs. low interaction state across each ICP pair in relation to progression-free survival (PFS). The forest plot presents results from the primary male cohort (brown bars, n=34) alongside the sensitivity analysis (green bars, n=46). An HR <1 indicates an association between higher ICP interaction and improved PFS; HR >1 indicates that higher interaction is associated with worse PFS. **(D)** Bivariate Cox proportional hazards regression analysis for high vs. low PD-1/PD-L1 interaction in relation to PFS using sex as covariable to estimate the confounding effect of sex on the association between PD-1/PD-L1 interaction and PFS. The forest plot presents the HR with 95% CI for PD-1/PD-L1 interaction status from the primary male cohort (brown bar, n=34) as a comparison alongside the HR for PD-1/PD-L1 interaction status corrected for sex (black bar, n=46) and HR for male vs female (grey bar; n=46). The arrow indicates the direction of effect associated with improved PFS.

### Association of immune checkpoint engagement with response to atezolizumab

To determine whether ICP interaction states in the urothelial cohort could inform on patient response to the atezolizumab treatment regimen, we first characterised the clinical heterogeneity of the cohort, as imbalances in baseline parameters could confound subsequent biomarker analyses. Clinical characteristics were examined across all patients (n=46), male (n=34) and female (n=12) separately (Table 1). This stratification was motivated by the fact that a patient’s gender is an established determinant of immune checkpoint inhibitor responses in urothelial cancer [16]. Urothelial carcinoma itself is a predominantly male disease, with an incidence ratio of approximately 3:1, and mechanisms of immune evasion may differ between sexes. Hormonal and immunological differences could therefore potentially act as confounding factors in biomarker-outcome associations.

The analysis of the clinical response revealed a marked imbalance in progression rates between sexes where 7 of 12 female patients (58.3%) experienced disease progression (PD) compared with 11 of 34 male patients (32.4%). In addition, the overall response rate (ORR) difference between male and female (38.2% vs 16.7%) also suggested an imbalance in the response to atezolizumab. Furthermore, in a univariate Cox regression analysis to evaluate the impact of the clinical parameters on PFS (Supplementary Fig.2A), despite not reaching significance, the female group showed a trend towards having a worse PFS with a hazard ratio (HR) = 2.17 (95%CI 0.79 – 5.54; Log-rank *P* = 0.1), while other risk factors like, age, prior lines or tumour stage with a HR close to 1 and wide 95%CI did not present any major bias that could substantially influence the observed results. Altogether, given this clinical imbalance and the known influence of sex hormones on immune regulation described in the literature, female patients were excluded from the primary survival analysis to reduce the biological confounding which may hinder a potential effect of ICP interaction status on PFS. Therefore, the male only cohort was analysed as the primary dataset, with the full cohort examined in a sensitivity analysis to test the robustness of the primary analysis using the same cutoffs.

The association between the three ICP interaction scores and overall best response was examined (Supplementary Fig. 2B). The ICP interaction scores distribution was compared in two groups: responders (CR+PR) and non-responders (SD+PD). The median distribution was assessed for each ICP (red bar). No significant difference in ICP interaction status could be observed between the responder versus the non-responders despite a trend towards higher interaction scores in PD-1/PD-L1 associated with the responders. This trend observed in the primary analysis (n=34) was retained in the full cohort (n=46).

To further assess whether of ICP interaction status could identify better responders to atezolizumab, we performed a univariate Cox proportional hazards regression evaluating the association between each checkpoint interaction levels (High versus Low interaction) and progression-free survival (PFS). Optimal stratification cutoffs for each interaction were determined independently (Supplementary table 1) using Cutoff Finder [17], an established online tool for cutoff selection in survival analyses (see Materials and Methods). The same cutoffs were applied without modification to both the primary male cohort (n=34, brown bars) and to the sensitivity analysis on the full cohort (n=46, green bars; Fig. 2C) ensuring methodological consistency.

In the primary cohort (Fig. 2C, red bar), higher PD-1/PD-L1 interaction was significantly associated with longer PFS compared to low interaction states (HR=0.26, 95% CI 0.06– 0.91). These data indicate that these atezolizumab treated patients with high PD-1/PD-L1 interaction had approximately 4-fold lower risk of progression compared to those with low interaction. Conversely, high CTLA-4/CD80 interaction was associated with shorter PFS at borderline significance (HR=3.43, 95% CI 1.00–15.65), suggesting that elevated CTLA-4/CD80 engagement may reflect a more atezolizumab resistant immune suppressive state associated with poorer outcomes. TIGIT/CD155 interaction did not reach significance in the primary analysis (HR=0.45, 95% CI 0.13–1.56).

In the sensitivity analysis (n=46), all three associations were directionally consistent with the primary results but did not reach statistical significance: PD-1/PD-L1 (HR=0.57, 95% CI 0.22–1.45), CTLA-4/CD80 (HR=2.42, 95% CI 0.94–6.94), and TIGIT/CD155 (HR=0.54, 95% CI 0.21–1.46). The attenuation of effect size and loss of significance in the full cohort is consistent with the known biological and clinical heterogeneity introduced by including female patients. Taken together, these data support PD-1/PD-L1 interaction as the strongest independent predictor of PFS in this cohort, with a suggestive trend for CTLA-4/CD80 in the opposite direction.

To confirm the confounding effect of sex on the PD-1/PD-L1 interaction–PFS association, a bivariate Cox regression analysis was performed in the full cohort (n=46), incorporating sex as a covariate (Fig. 2D). In the univariate analysis of the full cohort, the protective effect of high PD-1/PD-L1 interaction was attenuated compared to the male-only cohort (HR=0.57, 95% CI 0.22–1.45), consistent with the diluting influence of sex-related heterogeneity. Upon adjustment for sex in the bivariate model, the PD-1/PD-L1 FRET interaction HR strengthened towards the male-only estimate (HR=0.46, 95% CI 0.17– 1.21), while female sex independently emerged as a risk factor for disease progression (HR=2.66, 95% CI 0.94–7.04). The convergence of the bivariate-adjusted HR in the full cohort towards the male-only univariate (HR=0.26) demonstrates that sex was partially masking the PD-1/PD-L1 prognostic signal in the unajusted full cohort analysis, and validates the use of the male cohort as the primary analysis population to reveal the unconfounded biomarker-outcome association.

### High PD-1/PD-L1 interaction state is associated with improved PFS while High CTLA-4/CD80 associates with poorer PFS in Kaplan-Meier analyses

Kaplan–Meier analysis revealed significantly prolonged progression-free survival (PFS) in patients with high PD-1/PD-L1 interaction (*P* = 0.03; Fig. 3A). Median PFS was not reached in the High group (median undefined) versus 5.2 months in the Low group. With a hazard ratio of 0.26 it indicated about 74% reduction in the hazard of progression for patients with high PD-1/PD-L1. These findings suggest that patients with greater engagement of PD-1 and PD-L1 at baseline may derive preferential benefit from atezolizumab-mediated PD-L1 blockade.

**Figure 3.**
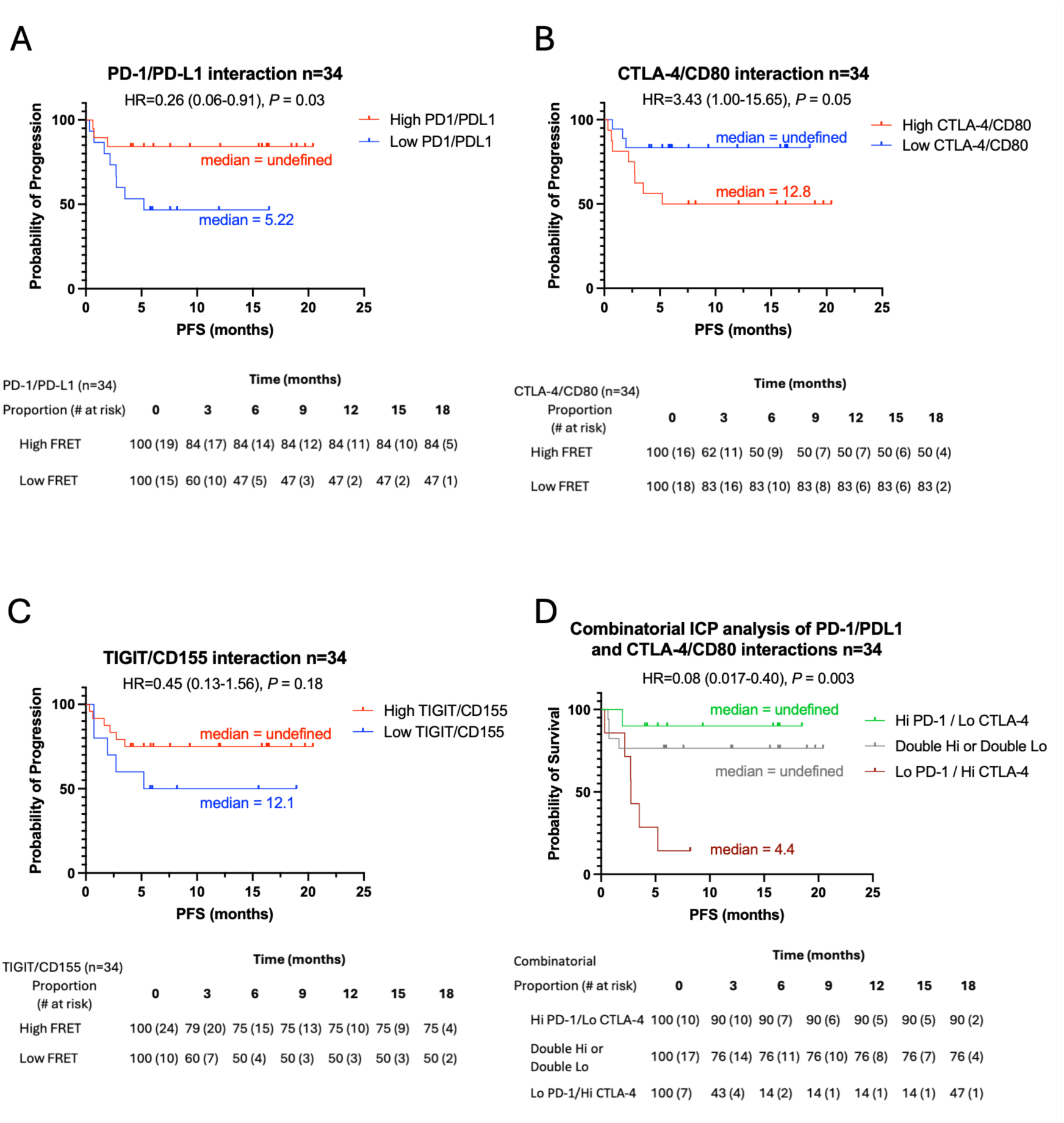
Progression-free survival stratified by immune checkpoint interaction in male urothelial carcinoma patients (n=34). (A-C) Kaplan–Meier analysis for progression-free survival stratified by ICP interaction levels for the three immune checkpoint pairs as indicated in male patients (n=34). Patients were classified as High (red) or Low (blue) interaction states using optimal cutoff values derived from Cutoff Finder (Materials and Methods). Statistical comparison was performed using the log-rank test; hazard ratios (HR) and 95% confidence intervals were estimated by Cox proportional hazards regression. **(D)** Kaplan-Meier analysis for PFS stratified by combined ICP interaction state: High PD-1/PD-L1 and Low CTLA-4/CD80 (green curve), Double High or Double Low for each checkpoint (grey curve), and Low PD-1/PD-L1 and High CTLA-4/CD80 (brown curve). HR = 0.08 and 95% confidence interval are calculated for the combined High PD-1/PD-L1 and Low CTLA-4/CD80 versus Low PD-1/PD-L1 and High CTLA-4/CD80 ; *P* = 0.003 shows the highly significant difference in the median PFS. For each Kaplan-Meier analysis the proportion and patients at risk over time is shown below the graphs.

Stratification by CTLA-4/CD80 interaction level however revealed the opposite directional association: patients with high CTLA-4/CD80 interaction exhibited shorter PFS (*P* = 0.053; Fig. 3B). Median PFS was 5.2 months in the High group versus not reached in the Low group. Although the P value did not reach the conventional <0.05 threshold, the effect size was substantial and suggested that a high CTLA-4 checkpoint engagement at baseline identifies patients who respond poorly to atezolizumab treatment. For the TIGIT/CD155 interaction, Kaplan–Meier analysis showed a trend towards improved PFS in patients with high TIGIT/CD155 proximity that did not reach statistical significance (*P =* 0.18; Fig. 3C). Median PFS was not reached in the High group versus 5.2 months in the Low group. It is worth noting that the limited statistical power in the Low group (n=10) may have constrained the ability to detect a true association. Given the established role of CD155 overexpression in urothelial carcinoma and its correlation with T cell exhaustion, the observed trend may warrant evaluation in a larger cohort. For each analysis a patients at risk table is presented under the curves.

To assess the robustness of the primary findings, Kaplan-Meier analyses were repeated in the sensitivity analysis including female patients (n=46), using the same cutoffs derived from the primary male cohort (Supplementary Fig. 3). All three directional associations were preserved: high PD-1/PD-L1 FRET remained associated with improved PFS (HR=0.57, *P* = 0.223), high CTLA-4/CD80 FRET with shorter PFS (HR=2.41, *P* = 0.069), and high TIGIT/CD155 FRET with a trend towards improved PFS (HR=0.54, *P* = 0.193). The proportion and patients at risk over time for these three groups are shown below the curves. None of the associations reached statistical significance in the full cohort, consistent with the hypothesis that inclusion of female patients, who had a higher baseline progression rate, introduced confounding that attenuated effect sizes. The direction and magnitude of effect estimates confirmed the directional consistency across both analyses, supporting the validity of the primary findings.

### Combinatorial PD-1/PD-L1 and CTLA-4/CD80 analysis predict better response to atezolizumab

Given that high PD-1/PD-L1 FRET interaction was associated with longer PFS and high CTLA-4/CD80 interaction with shorter PFS, we next asked whether combining these two biomarkers could further stratify patient outcomes (Fig. 3D). Patients in the primary male cohort (n=34) were classified according to their PD-1/PD-L1 and CTLA-4/CD80 interaction levels using the calculated cutoffs (0.501 and 5.578, respectively), yielding 3 groups: High PD-1/PD-L1 / Low CTLA-4/CD80 (n=10), double high PD-1/PD-L1 / CTLA-4/CD80 and double Low PD-1/PD-L1 / CTLA-4/CD80 (n=17 patients) were merged into one category, and Low PD-1/PD-L1 / High CTLA-4/CD80 (n=7). Kaplan-Meier analysis of these three groups revealed a significant separation in PFS (*P* = 0.012), with the High PD-1/PD-L1 / Low CTLA-4/CD80 group showing the most favourable outcomes and the Low PD-1/PD-L1 / High CTLA-4/CD80 group the poorest. This suggests that concurrent assessment of both checkpoint interactions provides stronger prognostic information than either biomarker alone. The proportion and patients at risk over time for these three groups are shown below the curves.

Together, these data indicate that the PD-1/PD-L1 and CTLA-4/CD80 interaction scores provide complementary and partially opposing prognostic information in urothelial carcinoma, and that their combined evaluation refines patient stratification beyond what either biomarker achieves individually.

## Discussion

Despite the transforming impact of immunotherapy directed at the PD-1/PD-L1 axis, many patients still do not derive durable benefit [5]. PD-L1 expression based on immunohistochemistry (IHC) measurements has been inconsistently predictive across various trials, suggesting that this biomarker may not be clinically useful in urothelial cancer [12, 13]. This inconsistency, attributed to the inherent limitations of quantification based on IHC, may also reflect the fundamental gap between protein amount and functional receptor engagement.

In this study, we used multiplexed FRET-FLIM to quantify immune checkpoint PD-1/PD-L1, CTLA-4/CD80 and TIGIT/CD155 interaction levels at baseline in advanced urothelial carcinoma FFPE TMA samples of patients treated with atezolizumab (n=46). Assessment of immune checkpoint interaction levels extends beyond the simple measurement of checkpoint protein expression and more faithfully reflects the functional targets of ICP therapies. Furthermore, the simultaneous quantification of the three ICP interactions in the same tumour samples further provided the opportunity to obtain a more comprehensive immunosuppressive profile for each patient.

The analysis of the three ICP interaction scores distributions (Violin plots) reflected intra-and inter-patient heterogeneity with variability in the extent of co-regulations or exclusivity of these three interactions depending on the patients. A pairwise Pearson’s r correlation analysis of the full cohort revealed that the three ICP were largely independently engaged across the tumours, suggesting tumour resistance to atezolizumab in this cohort was perhaps driven through distinctive immunosuppressive mechanisms.

Survival analyses were performed to determine whether any of the ICP interaction states could be associated with patients’ PFS. However, prior to performing the analyses, we evaluated whether clinical heterogeneity of the cohort may confound potential associations due to imbalances in baseline parameters.

Emerging evidence has shown that patient’s gender was an established determinant of ICI responses in urothelial cancer [16]. Recent genomic studies have identified sex-specific somatic alterations, including differences in RB1 and androgen receptor pathway alterations, that were associated with differential survival and distinct pattern of immune infiltration in UC [18]. Moreover, studies based on B-cell gene signatures have shown that immune configurations associated with ICI benefit in men were not predictive in women, suggesting that sex-specific differences in TME influenced therapeutic response [19]. Together it implied that sex may act as confounding factor in biomarker-outcome associations in this study. In accordance with these prior observations, the analysis of the clinical parameters of the cohort revealed that female patients had a substantially higher baseline rate of disease progression on atezolizumab treatment (58.3% versus 32.4% in males) with an ORR approximately half in females (16.7% versus 38.2%) albeit not quite reaching significance. Since these results directionally aligned with the sex-specific disparities described in the literature, females were excluded from the primary biomarker analysis (male cohort; n=34) to reduce biological confounding which may obscure the ICP interaction-survival associations, and then, reintroduced in a sensitivity analysis on the full cohort (n=46).

For each ICP, patients in the primary and sensitivity cohorts were divided into high and low interaction groups. The cutoffs for each ICP were derived from the male cohort using the web-based tool Cutoff finder (see Materials and Methods). Cox proportional hazard analysis showed that higher PD-1/PD-L1 interaction was significantly associated with better PFS in the male cohort (HR=0.26 95%CI 0.06–0.91; n=34). These results were concordant with prior findings where PD-1/PD-L1 interaction predicted response to anti-PD-1 axis blockade in non-small cell lung cancer [14, 20]. The consistent directional effect of this ICP interaction in the sensitivity analysis of the full cohort (HR=0.57 95%CI 0.22–1.45; n=46), further supported the association between higher PD-1/PD-L1 interaction and better PFS seen in the primary analysis, while the attenuated effect sizes and loss of statistical significance supported the female as a biologically distinct confounding subgroup in the analysis.

To further assess the impact of the female group on the association between high PD-1/PD-L1 interaction and better PFS, a bivariate Cox regression analysis was performed in the full cohort using sex as a covariate. The hazard ratio for the association of high PD-1/PD-L1 interaction and PFS corrected for the effect of sex (HR=0.46 95%CI 0.17–1.21; n=46, black bar) also revealed a consistent directional effect of the ICP interaction. The introduction of sex as a covariate led to a reduction in the HR as compared to the sensitivity analysis of the full cohort without correction (HR=0.57; n=46, green bar) aligning more closely to the HR of the male only cohort (HR=0.26; n=34, brown bar). This again supported the notion of bias introduced by the female group in PD-1/PD-L1 interaction outcome in the full cohort.

Kaplan-Meier survival analyses in the male cohort revealed that high PD-1/PD-L1 interaction was significantly associated with longer PFS (HR=0.26, Log-rank *P* = 0.03), corroborating the Cox proportional hazards analysis. The high-interaction group did not reach median PFS over the observation period of approx. 20 months, while the lower group displayed a median PFS of 5.22 months. Conversely, high CTLA-4/CD80 interaction at baseline was associated with shorter PFS (HR = 3.43, Log-rank *P* = 0.05). These data suggested that high CTLA-4/CD80 interaction status in urothelial tumours identified a patient subgroup in whom PD-L1 blockade appeared to be insufficient to drive a response.

Performing a combinatorial analysis integrating PD-1/PD-L1 and CTLA-4/CD80 interactions produced the most discriminatory result (HR=0.08, 95% CI 0.017–0.40, Log-rank *P* = 0.003). Patients with high PD-1/PD-L1 and low CTLA-4/CD80 interactions had substantially prolonged PFS with median not reached, while those with the inverse profile (low PD-1/PD-L1, high CTLA-4/CD80) experienced rapid progression (median PFS=4.4 months). These results were consistent with the fact that tumours having high PD-1/PD-L1 engagement would be more likely sensitive to PD-L1 blockade (atezolizumab), while harbouring CTLA-4/CD80 engagement, may evade anti-PD-L1 therapy through this alternative immunosuppressive pathway. Consequently, tumours displaying double high PD-1/PD-L1 and CTLA-4/CD80 interactions may represent candidates for combinatorial PD-1 + CTLA-4 blockade strategies.

The analysis of TIGIT/CD155 interaction showed a trend towards favourable PFS in the high-interaction group (HR=0.45, Log-rank *P* = 0.18), with a median not reached versus 12.1 months. Despite not reaching statistical significance likely due to the small sample size, the marked difference in the median PFS, would justify exploration in larger cohorts.

The association of the three ICP interaction status with PFS was then explored in the sensitivity analysis. In the expanded cohort including female patients (n=46), the direction of all associations was preserved but showed attenuation of statistical significance for PD-1/PD-L1 (HR=0.57, *P* = 0.22) and CTLA-4/CD80 (HR=2.42, *P* = 0.07), TIGIT/CD155 (HR=0.54, *P* = 0.19). These results were consistent with the described sex-related differences in treatment response in urothelial carcinoma.

Altogether these findings suggest that high PD-1/PD-L1 interaction status may be useful as a predictive biomarker of response to atezolizumab while, the engagement of CTLA-4/CD80 interaction at baseline associated with shorter PFS may represent an adverse prognostic factor. While PD-1/PD-L1 engagement primarily attenuates effector T cell function at the tumour site, the CTLA-4/CD80 axis operates predominantly at the priming phase, where CTLA-4 expressed on Tregs competes with CD28 for binding to CD80 and CD86 on APCs. High-affinity CTLA-4/CD80 engagement at baseline would compromise effector T cell priming, thereby enforcing an immunosuppressive TME which could not be overcome by PD-L1 blockade alone regardless of the degree of PD-1/PD-L1 interaction on effector cells. This aligns with growing evidence that dual CTLA-4 and PD-1/PD-L1 blockade can act synergistically in urothelial carcinoma as demonstrated in the CheckMate032 trial [7]. Hence patients with high CTLA-4/CD80 engagement may be precisely those who would benefit most from combination anti-CTLA-4/anti-PD-L1 therapy, while they are currently treated with monotherapy. Prospective validation of CTLA-4/CD80 interaction as a selection biomarker in addition to PD-1/PD-L1 interaction for combination regimens would therefore merit close attention.

The TIGIT/CD155 axis may also represent a valuable dimension within a multiplex profiling strategy. CD155 is overexpressed in urothelial carcinoma and elevated TIGIT correlates with worse outcomes in muscle-invasive bladder cancer [9]. Therefore, in this study, the potential positive association between high TIGIT/CD155 interaction and benefit to atezolizumab, an intervention targeting a distinct checkpoint axis, could reflect an immunologically active TME. In this environment, effector T cells could be present but suppressed, thereby providing a highly favourable landscape for response to atezolizumab. In the context of recent clinical setbacks for anti-TIGIT agents, including the discontinuation of tiragolumab across multiple phase II/III trials and its failure in combination with atezolizumab, as well as the absence of validated predictive biomarkers for ongoing neoadjuvant anti-TIGIT strategies in UC (NCT05394337) [21], quantification of TIGIT/CD155 engagement may help identify the patients most likely to benefit from this therapeutic class.

Several limitations of the present study, however, deserve acknowledgment. Although the cohort size is consistent with exploratory biomarker studies in this disease context, it limits statistical power. Furthermore, its single-centre nature may introduce inherent selection biases that must be considered when extrapolating these findings to broader patient populations. Concerning the analyses of the cohort, the biomarker thresholds were determined using Cutoff Finder, a dataset-driven tool that may introduce risks of overfitting and optimization bias. However, alternative approaches such as median splits or quartile-based thresholds carry analogous limitations in small samples without providing a biological foundation. Consequently, prospective validation in an independent cohort with identified fixed cutoffs will be required before these ICP interactions can be considered clinically actionable.

In conclusion, this study suggests that although the evaluation of these findings in a larger cohort will be critical, functional multiplexed FRET-FLIM profiling of immune checkpoint interactions allowing simultaneous quantification of multiple checkpoint axes in standard FFPE material, may offer a clinically relevant biomarker strategy for treatment selection in urothelial carcinoma. Conceptually analogous to the combinatorial scoring approaches that have gained traction in other cancer types, such as combined TMB and PD-L1 assessment, FRET interaction scores derived from the same tissue in a single platform, may provide functional actionable cutoffs for identifying the optimal therapeutic regimen. As combination immunotherapy reshapes the treatment landscape of advanced urothelial carcinoma, the unmet need for biomarkers that reflect the functional immune state of individual tumours rather than the presence of a single protein will become increasingly acute, and the present findings provide a rationale for incorporating this approach in future clinical studies.

## Authors contribution

All authors listed have made a substantial contribution to the work and approved it for publication L.C. and C.C.N. designed and performed the QF-Pro experiments on FFPE samples. J.G., M.R. and J.M. were responsible for technical implementation of QF-Pro. C.E.deA. and S.M.A. carried out patient recruitment and provided the FFPE tumour samples and associated clinical data. J.A., B.B., J.M.G., K.O’S. and F.A. were involved in the development and maintenance of the QF-Pro FLIM platform software and hardware. V.C. performed the statistical analyses. C.E.deA. and S.M.A. and V.C. were involved in the critical discussion of the clinical results. V.C. wrote the manuscript. P.J.P., J.G. and J.M. were involved in the critical reading of the manuscript. All authors read and approved the final manuscript.

## Data Availability

All data produced in the present study are available upon reasonable request to the authors

## Acknowledgments

This work was supported by the Spanish Ministry of Science and Innovation and the State Research Agency (MCIN/AEI/10.13039/501100011033) under the project PREDICTEAM (Grant No. CPP2021-008390).

## Ethics statement

The study was conducted accordingly with the International Conference on Harmonization (ICH) Good Clinical Practice (GCP) guidelines, as they apply to observational research, and to the ethical principles outlined in the and European Union Directive 2001/20/EC and in the Declaration of Helsinki 2013, including all patient privacy requirements, which will also comply with European and Spanish regulations. All samples were used after approval from the Hospital Universitario Parc Taulí de Sabadell Human Research Committee (Protocol number ML40792/ROC-URO-2019–01) and all patients gave their written informed consent.

## Data availability statement

The data that support the findings of this study are available from the corresponding author upon reasonable request.

## Materials and methods

### Primary antibodies

PD-1 mouse mAb and PD-L1 rabbit mAb were provided by Abcam. TIGIT mouse mAb was from Promega, CD155 rabbit mAb was from Novus Biologicals. CTLA-4 mouse mAb was from Antibodies online and CD80 rabbit mAb was provided by MyBioSource.

### Study cohort

Retrospective observational, multicentre RWE study of patients presenting locally advanced or metastatic urothelial carcinoma who had progressed during or following a platinum-containing regimen and had received atezolizumab (anti-PD-L1) under a compassionate use program (n=109), described in de Andrea *et al.* [15]. Of these, 46 patients archival formalin-fixed, paraffin-embedded (FFPE) tumour tissue was available for biomarker analysis. FFPE were collected prior to treatment and presented in tissue microarrays (TMA). Samples were reviewed by two board-certified pathologists (C.E. A. and M.A.M.) using haematoxylin and eosin (HCE) stained slides. Tumour presence was confirmed by morphology. TMAs were constructed using standard procedures where 1,5 mm cores were obtained from the donor paraffin blocks using a needle and transferred to the recipient array block. For better representation, two cores from different areas of the tumours were included in the TMAs.

Baseline clinical characteristics including age, sex, disease setting (primary vs. metastatic), number of prior lines of therapy, and best response to atezolizumab (assessed per RECIST v1.1) were collected.

### QF-Pro (Quantifying Function in Proteins) Assay

Before incubation with QF-Pro reagents, pre-treatment processes of deparaffination, rehydration and epitope retrieval were performed using a Dako PT-Link. Samples were then treated following the QF-Pro labelling protocol previously described in Cacho-Navas *et al.* [22]. Briefly, samples were incubated with endogenous peroxidase suppressor and then with blocking buffer to prevent non-specific signal. Samples were incubated with primary antibodies overnight at 4°C and then with donor and acceptor secondary reagents at room temperature (RT) for two hours. Amplification reagent was applied to Donor-Acceptor slides for 20 min and mounting medium was applied. Slides were stored until ready for fluorescence lifetime imaging microscopy analysis.

### Multiplexed QF-Pro assay

Three immune checkpoint protein (ICP) pairs were interrogated in three sequential staining rounds on the same tissue section: (i) PD-1/PD-L1 (Round 1), (ii) CTLA-4/CD80 (Round 2), and (iii) TIGIT/CD155 (Round 3). Each round involved dual-labelling of the receptor–ligand pair with primary mAbs followed by spectrally compatible secondary antibodies (anti mouse F(ab’)2-ATTO488) donor and ALEXAFluor594-tyramide acceptor amplification using anti-rabbit F(ab’)2-HRP. The fluorescent secondary reagents were modified to allow the removal of both fluorophores ATTO488 and ALEXAFluor594 from previous rounds of labelling before to proceeding to new rounds. Therefore, a series of custom developed TCEP cleavable disulphide fluorescent compounds (cleavable Tyramide-AlexaFluor594 and cleavable ATTO488) were synthesized by Celtarys Research S.L. (Santiago de Compostela, Spain) and were used instead of the QF-Pro classical fluorescent secondary reagents. The conjugation of the cleavable ATTO488 to the secondary F(ab’)2 was performed by Hypermol EK (Bielefeld, Germany).

### QF-Pro FRET-FLIM platform

Protein–protein interactions between immune checkpoint receptor–ligand pairs were quantified using the QF-Pro platform Violet 3.0. (HAWK Biosystems), an amplified FRET-FLIM system designed for multiplexed analysis of FFPE tissue. Fluorescence lifetime imaging microscopy (FLIM) and QF-Pro software automatically measures the fluorescence decay of the lifetime (τ) of the donor fluorophore, which decreases (τDA) when Förster resonance energy transfer (FRET) occurs due to close proximity (<10 nm) between donor-and acceptor-labelled proteins. The interaction states (FRET efficiency (Eff %) values or QF-Pro scores) are calculated as followed:

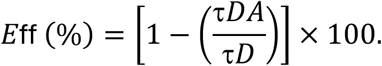

### Statistical analysis

Statistical analyses and data visualisations were performed in GraphPad Prism (v10). All two-tailed p-values <0.05 were considered statistically significant. Trend-level associations (0.05 ≤ p <0.10) are reported as such. Kaplan-Meier survival curves were generated and compared using the log-rank test. For the forest plot analyses univariate Cox proportional hazards regression was used to estimate hazard ratios (HR) and 95% confidence intervals for each ICP biomarker (high vs. low FRET interaction) and for clinical covariates (disease setting, sex, age, prior lines of therapy). Statistical significance between groups was assessed by the log-rank test. A dotted line referenced HR = 1 indicates the null effect.

Pairwise correlations between continuous FRET interaction scores for the three ICP pairs were assessed using Pearson’s correlation coefficient r. Potential outliers were identified using two independent methods applied simultaneously: Z-score (|z|>2.5) and Tukey’s interquartile range fences (values beyond Q1−1.5×IQR or Q3+1.5×IQR). Only data points flagged by both methods were considered outliers for exclusion; two outliers meeting both criteria were removed from the correlation analysis. Correlation matrix was exported directly from Prism 10 for presentation. Statistical significance was defined as *P* < 0.05.

For Combinatorial biomarker analysis, the patients were classified into four groups based on their PD-1/PD-L1 and CTLA-4/CD80 FRET interaction status (high/low). Kaplan-Meier analysis was performed on high PD-1/PD-L1 / low CTLA-4/CD80 against dual-low or dual-high group and low PD-1/PD-L1 / high CTLA-4/CD80 remaining patients.

Violin plots were used to show the distribution of patients’ sample lifetime distribution. The outer boundaries present the kernel density estimation illustrating the probability density of lifetimes at different values. The median is represented for each condition with the whiskers extended to represent the data variability.

### Biomarker cutoff determination

Optimal FRET interaction score cutoffs for patients’ separation into high and low groups were determined using Cutoff Finder (https://molpath.charite.de/cutoff), an online tool that identifies the threshold maximising the separation of survival outcomes by log-rank statistic. Cutoff derivation was performed exclusively on the primary male cohort (n=34). The same cutoffs were applied without modification to the sensitivity analysis on the full cohort (n=46) to ensure methodological consistency and avoid data-driven bias.

## Supplementary figures

**Supplementary figure 1:**
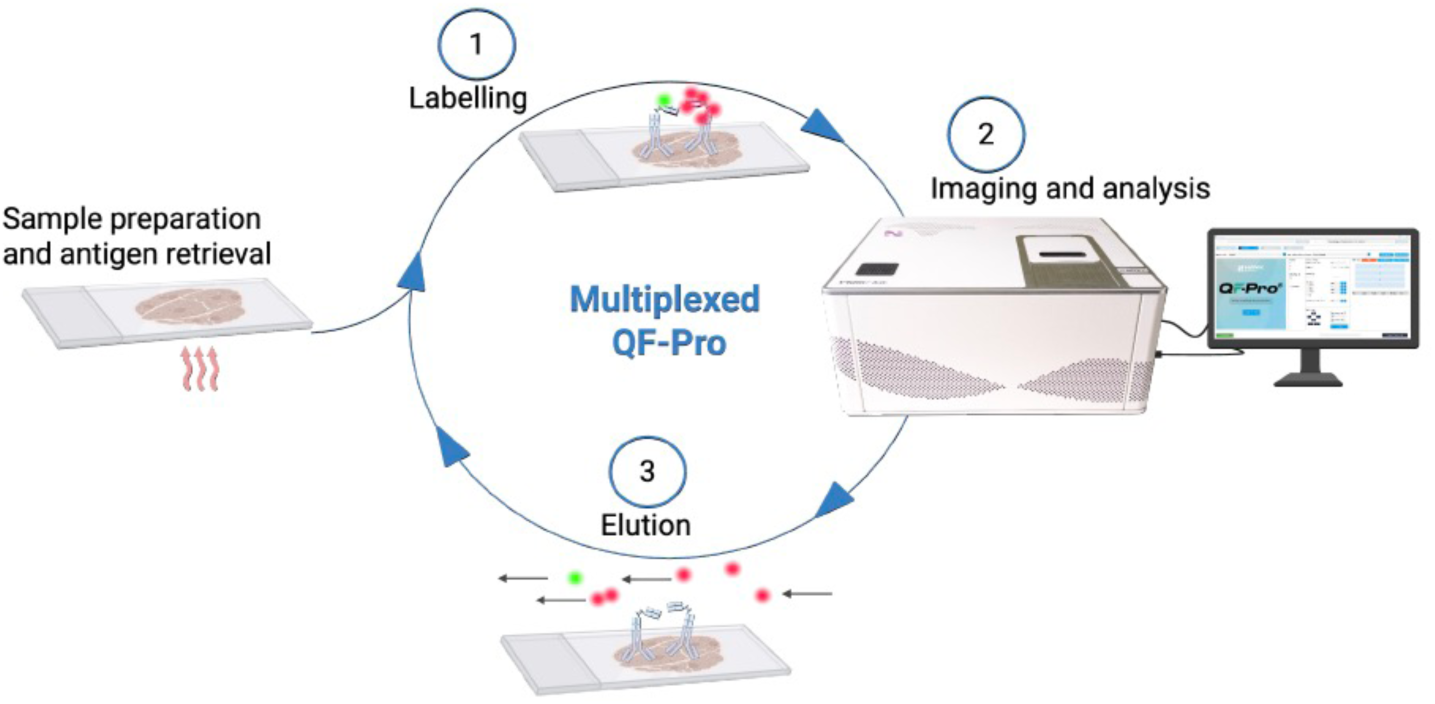
Tables and Supplementary tables **Multiplex QF-Pro workflow.** In step 1/ Following antigen retrieval, the FFPE TMAs were labelled with the first round of antibodies primary and secondary. Mouse and rabbit primary antibodies targeting respectively ICP receptor and ligand were detected using TCEP cleavable donor anti-mouse F(ab’)₂–S-S-ATTO488 (green) and anti-rabbit F(ab’)₂–HRP secondary antibodies. Signal amplification was achieved using TCEP-cleavable tyramide (acceptor, red), enabling detection of FRET. Step 2/ the samples were acquired in the FRET-FLIM violet 3.0 imaging platform and associated analysis software. Images of the ICP protein expression (fluorescence intensity) and interaction (QF-Pro maps) were obtained together with the quantification of the interactions (QF-Pro scores). Step 3/ for the following round of labelling the samples were treated with TCEP for 30 min at room temperature to cleave the fluorescent dyes of the first round. After PBS wash, a second round of labelling with the following protein pair can be performed as described in step 1.

**Supplementary Figure 2.**
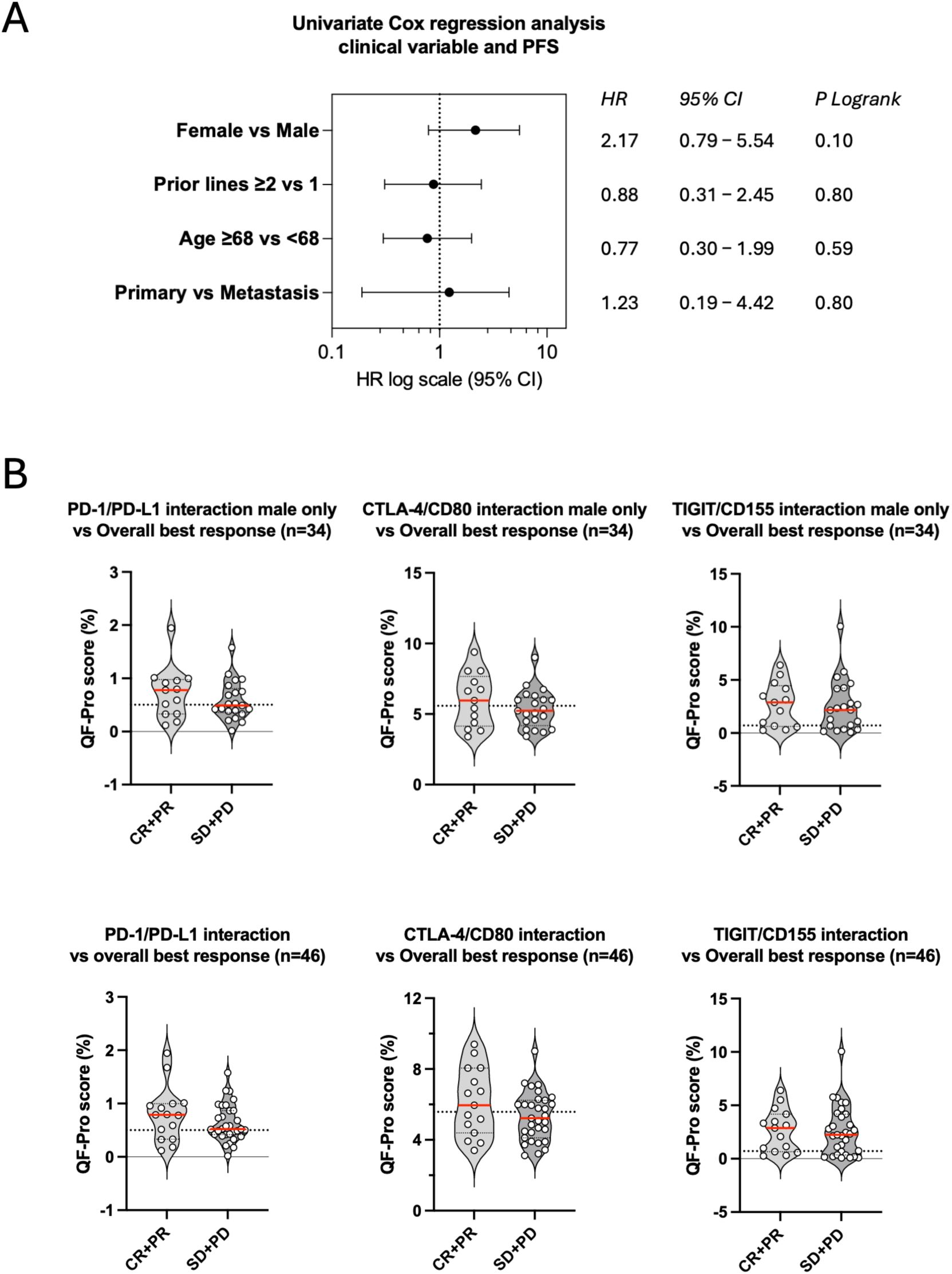
Full patients cohort (n=46) clinical characteristics and FRET-FLIM (QF-Pro ®) scores distribution. **(A)** Forest plot showing Hazard ratios (HRs) and 95% confidence interval (95%CI) derived from Cox regression analysis evaluating the association between the clinical parameters and PFS in the full patient cohort (n=46). **(B)** Violin plots showing the distribution of the QF-Pro scores for the three ICP in the male patient cohort (n=34) and the full cohort after reintroducing female patients (n=46). Interaction scores are shown separately for responders (CR+PR) and non-responders (SD+PD). Each dot represents an individual patient. Red horizontal lines indicate the median, and dotted lines denote the QF-Pro score cutoff for each ICP.

**Supplementary Figure 3.**
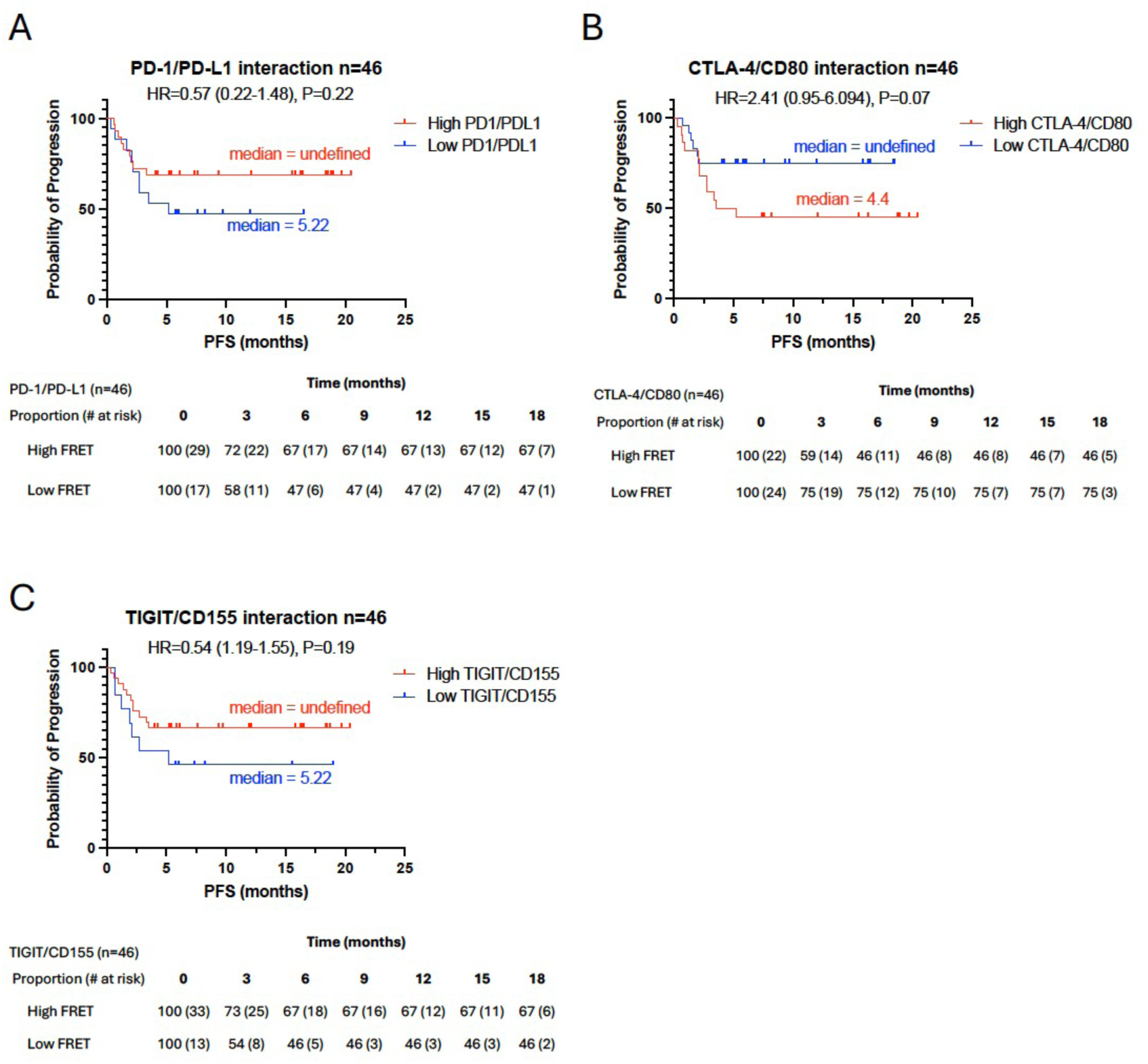
**Kaplan–Meier analysis of PFS stratified by ICP interaction levels in the full patients cohort (n=46)**. **(A-C)** Patients were classified into High (red) or Low (blue) interaction groups using cutoff values determined with Cutoff Finder (see Materials and Methods). Statistical significance was assessed using the log-rank test, and hazard ratios (HRs) with 95% confidence intervals (95%CI) were estimated by Cox proportional hazards regression. The number and proportion of patients at risk over time are provided in the tables below each graph.

## Tables and Supplementary tables

**Supplementary table 1.** : FRET-FLIM (QF-Pro) interaction scores ranges

|  | All patients<br>(n = 46) | Males<br>(n = 34) | Cutoff values for<br>n=34 |
| --- | --- | --- | --- |
| **PD-1/PD-L1** - median (range) | 0.60 (0.02–1.94) | 0.55 (0.02–1.94) | 0.501 |
| **CTLA-4/CD80** - median (range) | 5.44 (3.11–9.40) | 5.38 (3.41–9.40) | 5.578 |
| **TIGIT/CD155** - median (range) | 2.27 (0.08–10.06) | 2.16 (0.08–10.06) | 0.710 |

